# COVID 19 Knowledge, Attitude, and Practice of the Healthcare Providers in United Arab Emirates

**DOI:** 10.1101/2021.03.05.21252719

**Authors:** Aljazia Khalfan Alghfeli, Amal Abdul Rahim Al Zarouni, Hamda Musabbah Alremeithi, Roqayah Abdulla Almadhaani, Latifa Baynouna Alketbi

## Abstract

**Background:** Health care providers at increased risk of COVID-19 infection, inadequate knowledge and practice about COVID-19, and infection control may lead to increased risk of disease transmission. Early diagnosis and appropriate management of COVID 19 cases is important in preventing transmission and improving patient outcomes. The aim of this study was to assess the knowledge, altitude, and practice of healthcare providers in the United Arab Emirates toward COVID-19 and to examine its determinants.

**Method:** A cross-sectional study was conducted to assess knowledge, attitudes, and practice (KAP) of Healthcare providers regarding COVID-19. The study was conducted during the COVID-19 pandemic from of April 11 to July 23, using an online anonymous self-administered questionnaire.

**Results:** A total of 2371 healthcare providers responded to the survey. A total of 1091 worked in inpatient hospitals, 494 in primary health care, and 388 in emergency and ICU care. The overall performance score for all healthcare providers was as follows: 49.1%, poor score; 41.8 %, intermediate score; and 9.2%, good score with a mean result of 17.14. Factors leading to better overall performance scores were years of experience, pediatricians’ specialty, and specialist’s occupation. A total of 55.7% received good direct knowledge from all healthcare providers. In practice, 48% had good practices toward COVID-19. The overall attitude mean was 2.8, from a maximum score of 7, indicating a positive attitude toward COVID-19.

**Conclusions:** The study-demonstrated gaps in specific aspects of knowledge and practice that should be focused on in future education and HCP awareness. A structured training program targeting all HCPs is needed to have good clinical knowledge and practice about COVID-19.

## Introduction

The current emergence of COVID-19 creates a global health burden and public health crisis in many countries.^1^ COVID-19 was declared a global pandemic on March 11, 2020, by the World Health Organization (who).^2^ Covid-19 is responsible for mild to severe respiratory infections. In the UAE transition in the health care system, it happens to adapt to the pandemic public health needs. Governments focus on preventive measures and policies to control the COVID-19 pandemic that has impacted the economic, social, and mortality rates. Measures to prevent infectious disease transmission include frequent hand washing, avoiding direct contact with infected patients, vaccination and wearing masks.^3^ Health workers are currently making efforts to control further disease outbreaks caused by COVID-19.^1^ Health care providers are the frontline during COVID-19 pandemics; they are exposed to infection hazards in addition to psychological distress. Healthcare providers had a higher risk of exposure and acquired COVID-19 if personal protective measures were not used appropriately.^4^ Poor knowledge about communicable disease and infection control practices places them at increased risk. In 2002, during the SARS outbreak, approximately one-fifth of all cases were from health care workers. ^5^ A recent study conducted in the UAE demonstrated a gap in healthcare workers’ knowledge of COVID-19^6^. Health authorities have released COVID-19 national guidelines, much educational material, and online educational sessions for healthcare providers. Free access to online medical library resources was provided to healthcare workers with access to guidelines, policies, procedures, and recommendations about COVID-19. Healthcare providers can request articles about COVID-19. Risk assessment and periodic COVID-19 screening tests were performed for all healthcare providers in the United Arab Emirates. In addition, mental counseling, psychologist support, and helpline support were provided to the health care team. Healthcare workers play an important role in controlling infectious diseases. Infectious disease outbreaks is likely to impact the psychological health healthcare workers (HCWs) who are in the frontline in facing the pandemic ^7^.

The aim of this study was to assess the knowledge, attitude, and practice of healthcare providers in the United Arab Emirates toward COVID-19. In addition, to examining factors influencing their adherence to precautionary practice.

## Methods

### Study Design, Setting and Population

A cross-sectional study was conducted in the United Arab Emirates among healthcare providers from primary health care centers and hospitals in the private and public sectors. The study was conducted from during April 11 to July 23. The health care provider in Seha’s sample size was determined using a margin of error of 5%, and a confidence interval (CI) of 95%, Due to the distribution of the survey as online sampling, it was a convenient sample.

### Survey Design

Data were collected using an anonymous online self-administered questionnaire developed in English. The survey was designed to collect information about the demographic data of the participants (age, occupation, specialty, years of experience, city, and practice sitting). The survey was intended to collect information about healthcare providers’ knowledge, practice, and attitude about and toward COVID-19, information collected about basic knowledge on COVID-19, guidelines, mode of transmission, investigation, risk group, infection prevention, exposure risk assessment for healthcare workers, and action. Screening for depression and anxiety using PHQ9 and GAD 7 was performed. The questionnaire was developed by reviewing available questionnaires in the literature and the national, SEHA, CDC, and WHO guidelines. The survey included expert opinions and available guidance. The question design is a case-based scenario. The questionnaire was piloted among 20 healthcare providers.

The scoring system was developed by assigning one score to each correct answer. The survey included direct knowledge, practice, and attitude questions in base case seniors that measure knowledge, practice, and attitude. The overall performance of the participants was measured by considering the survey as a quiz-like assessment of real-life situations. The total score of overall performance was 40. Based on the 40 questions available in the survey. An overall performance score of more than 30 indicates good performance. An overall performance score of less than 30 indicates poor performance. Intermediate performance was assessed using a 20-30 performance score. There were 18 direct knowledge-based questions, seven attitude base questions, and nine direct practice base questions.

The knowledge section total score ranges from 0 to 18, and a cut-off level of ≤ 8 was set as poor knowledge and ≥ 9 for good knowledge. The attitude section’s total score ranged from 0 to 7. Higher values indicated a positive attitude. The practice items total score ranged from 0 to 9, and a score of <5 indicated poor practice toward precautionary measures of COVID-19.

The depression scoring system includes nine items. Each item had four responses. Each response had a certain score as follows: not at all=0, several days=1, more than half a day=2, nearly every day=3. The total scores for depression items ranged from 0 to 27, 0 to −4 indicated minimal depression, 5 to 9 indicated mild depression, 10 to14 indicated moderate depression, 15 to 19 indicated moderately severe depression, and 20 to 27 indicated severe depression.

The generalized anxiety disorder scoring system includes seven items. Each item has four responses, and each response has a certain score as follows: not at all=0, several days=1, more than half a day=2, nearly every day=3 Depression items’ total score ranged from 0 to 21. A total score of 0 to 4 indicated minimal anxiety, 5 to 9 indicated mild anxiety, 10 to 14 indicated moderate anxiety, and 15 to 21 indicated severe anxiety.

## Statistical Analysis

Descriptive statistics such as mean, standard deviation (SD), median, and interquartile range. (IQR) were computed for quantitative variables, and frequencies and percentages were calculated for categorical variables. All analyses were performed using SPSS 21 software program. A Chi-square test was conducted to determine the association between KAP and the independent categorical variable. The significance value of ≤ 0.5 was set at the significant level.

## Ethics and Confidentiality

All study participants were informed of the study. Online consent for participation was obtained before enrollment. This study was approved by the SEHA Ethics Committee. Institutional Review Board and conducted in accordance with the Ethics Committee guidelines The questionnaire was anonymous and did not include any identifiers or personal information of the participants. The confidentiality of the participants was maintained.

## Results

### Characteristics of the Study Population

A total of 2371 healthcare providers responded to the survey. The excellent response rate was achieved from April to July. 46% of them (1091) were working in inpatient hospitals, 20.8% (494) were working in primary health care and 16.4% (388) were working in emergency and ICU care settings. The majority of participants 1926 were from SEHA and only 225 were from outside SEHA. Participants had a mean age of 39.94 years and were mostly female (74 %). Nurses constituted 61.9 % of the respondents and 18.8% were physicians (consultants, specialists, residents), 13.9 % were technicians, and only 5.4% were pharmacists. Physicians who enrolled were from different specialties: family medicine, 7%; internal medicine, 8.6 %; obstetrics and gynecology, 7.6%; pediatrics, 9.7%; psychiatry, 1.6%; and surgeons, 8.8%. Most of the participants were from Abu Dhabi and Al Ain (64%, 26.5%), respectively, and 8.1% of participants were from the western region. The mean work experience was 14 years. The demographic characteristics of the study population are summarized in Table 1.

**Table 1.** Characteristics of the study population

| **Table 1. Characteristics of the study population** |  |  |
| --- | --- | --- |
| Age (years) | Mean age | 39.94 years |
| Sex % (N ) | Female | 74.0 (1754) |
|  | Male | 26.0 (617 ) |
| Occupation % ( N ) | Consultant | 3.9 ( 93 ) |
|  | Nurse | 61.9 (1467) |
|  | Pharmacist | 5.4 (129) |
|  | Resident | 3.8 (90 ) |
|  | Specialist | 11.1 (262) |
|  | Technician | 13.9 (330) |
| Specialty % ( N ) | Family medicine | 7.0 (166) |
|  | Internal Medicine | 8.6 (205 ) |
|  | Obstetrics and Gynecology | 7.6 (180 ) |
|  | Other | 56.6 (1343) |
|  | Pediatrics | 9.7 (230) |
|  | Psychiatry | 1.6 (39) |
|  | Surgical specialties | 8.8 (208) |
| Practice sitting % | Emergency /ICU | 16.4 |
|  | Inpatient hospital based | 46 |
|  | Other | 16.8 |
|  | Primary health care clinic | 20.8 |
| City % | Abu Dhabi | 64.0 |
|  | Alain | 26.5 |
|  | Other | 1.3 |
|  | Western Region | 8.1 |
| Experience | Years of work experience (mean) | 13.988 years |

### COVID-19 Overall Performance Score

The overall performance score among healthcare workers was divided into three groups based on their achievement from a total of possible 40 points if all were correct answers. Poor performance scores were given for those who achieved less than 20 out of 40. The intermediate performance scores ranged from 20 to 30. Good performance scores were given for those who scored above 30 points. Almost half of the participants (49.1%) were poor performers, 41.8 % had intermediate performance, and only 9.2% had a good performance score. The overall mean score was 17.14.

Of all factors studied, only years of experience, being in pediatricians, and specialist physicians’ positions showed significantly better overall performance scores than others (B=1.881, 1.968, 0.065 p =0.012, 0.013, 0.022, respectively). Figure 1 shows the overall performance scores among the specialties.

**Figure 2.**
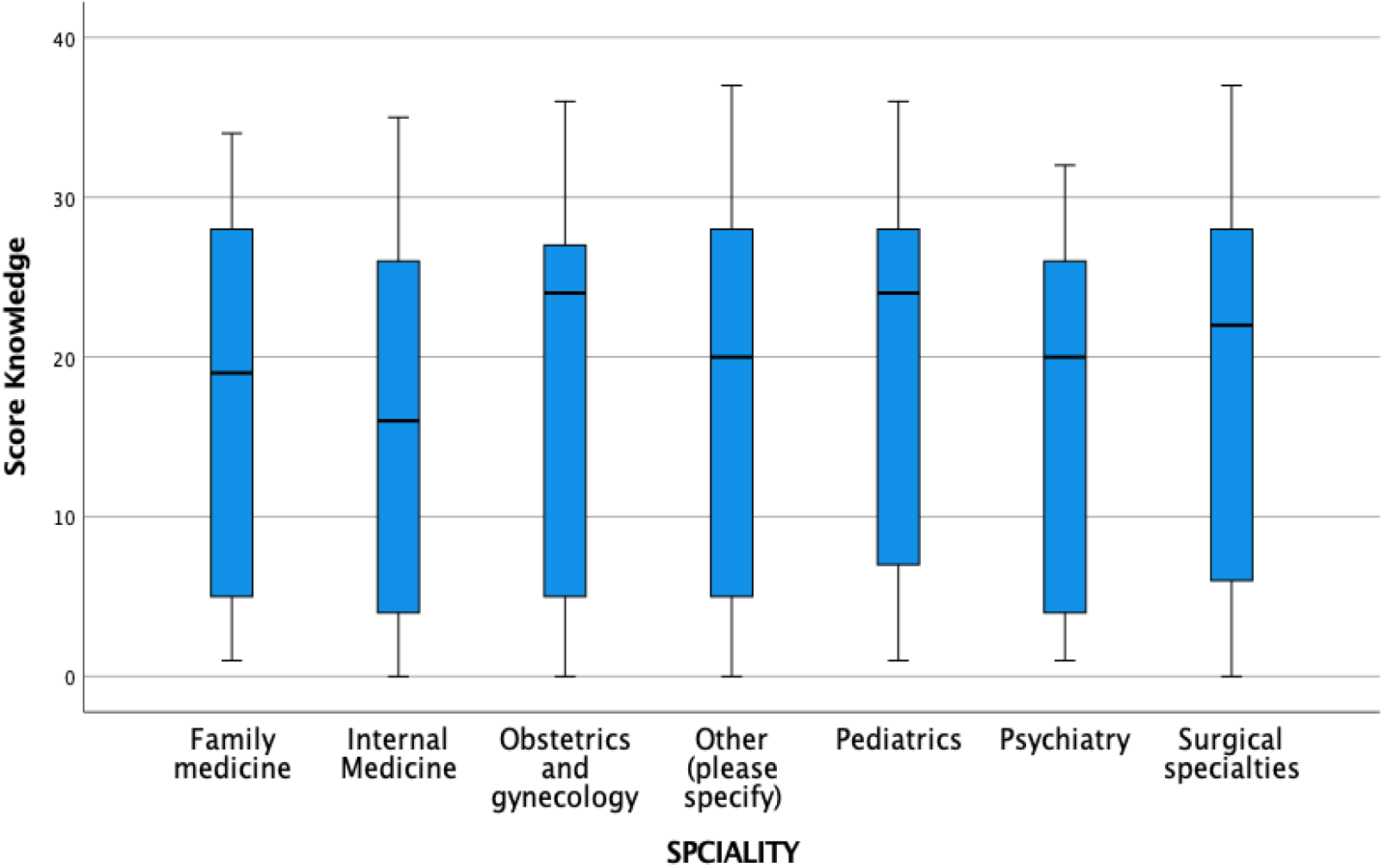
overall performance score distribution among specialty.

Regarding knowledge questions, only half of the respondents indicated that the mode of transmission of COVID-19 virus was by respiratory droplets, 55.5% (believed that transmission by contact with contaminated surfaces and 32.3% believed that COVID-19 virus transmission is via the oral route. Social distancing of 2 m or more was reported by 43.3% of healthcare providers as the correct distance to prevent transmission of COVID-19 virus, while 11.3% of all healthcare providers believed that 1 m or more was considered a safe social distance and 0.1% reported that they did not know the exact social distance.

More than half of healthcare providers reported the following as high-risk groups: age >60 years (55.5%), smokers (50%), diabetes mellitus (53.6%), hypertension (50.3%), patients with chemotherapy (55.1%), and patients with asthma and COPD (55.1%). Pregnant women, 48.2%, were also reported as a high-risk group.

Nasopharyngeal swab was reported by 39.7% to be recommended over the oropharyngeal swab for detecting COVID-19 virus;with regard to the PCR test sensitivity and specificity, 17.8% of all healthcare providers believe that the sensitivity of the nasopharyngeal swab is 90%, while 13.5% think that the sensitivity is 70 % and 8.3% think that the specificity of the nasopharyngeal swab is 70%.

Wearing surgical masks was perceived by 32.2% of all healthcare providers to be necessary for suspected COVID-19 infection patients, and 21% reported the need to wear surgical masks with patients who have only respiratory symptoms such as fever and cough. While 20% reported that only medical staff and caregivers in close contact with patients should wear surgical masks, and less than half of the healthcare providers (43.4%) reported that all communities should wear surgical masks. Table (2)

**Table 2:**
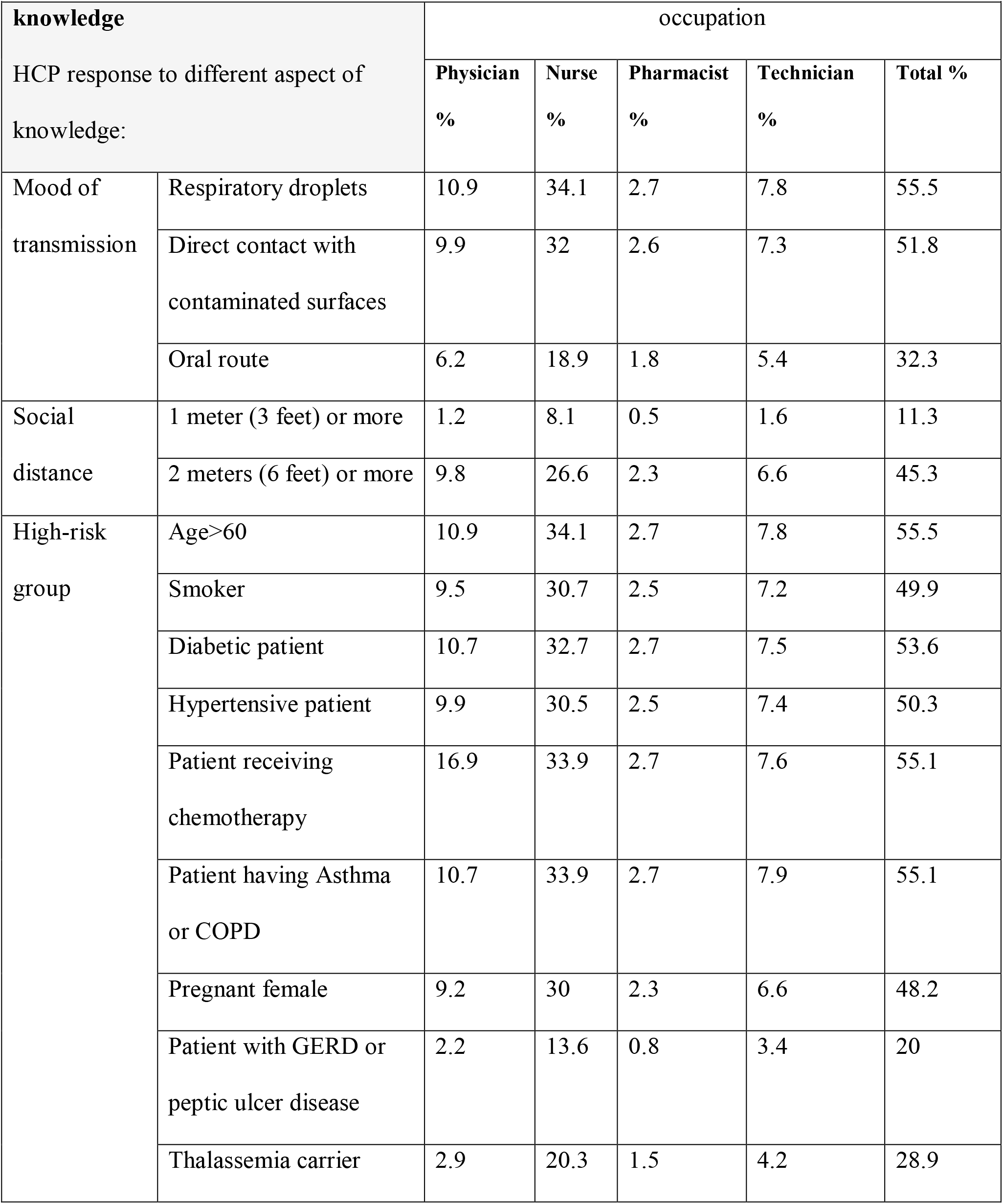
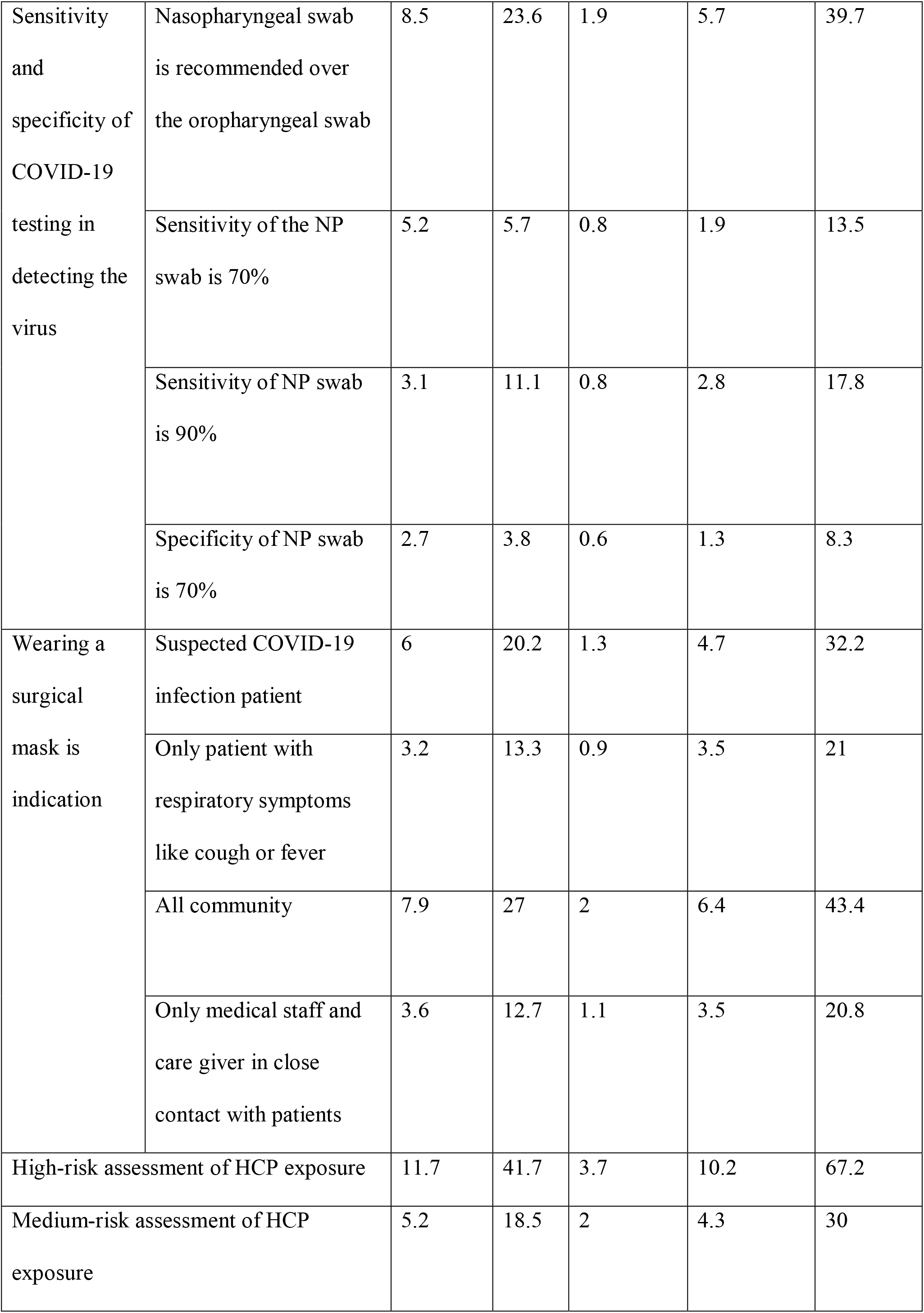
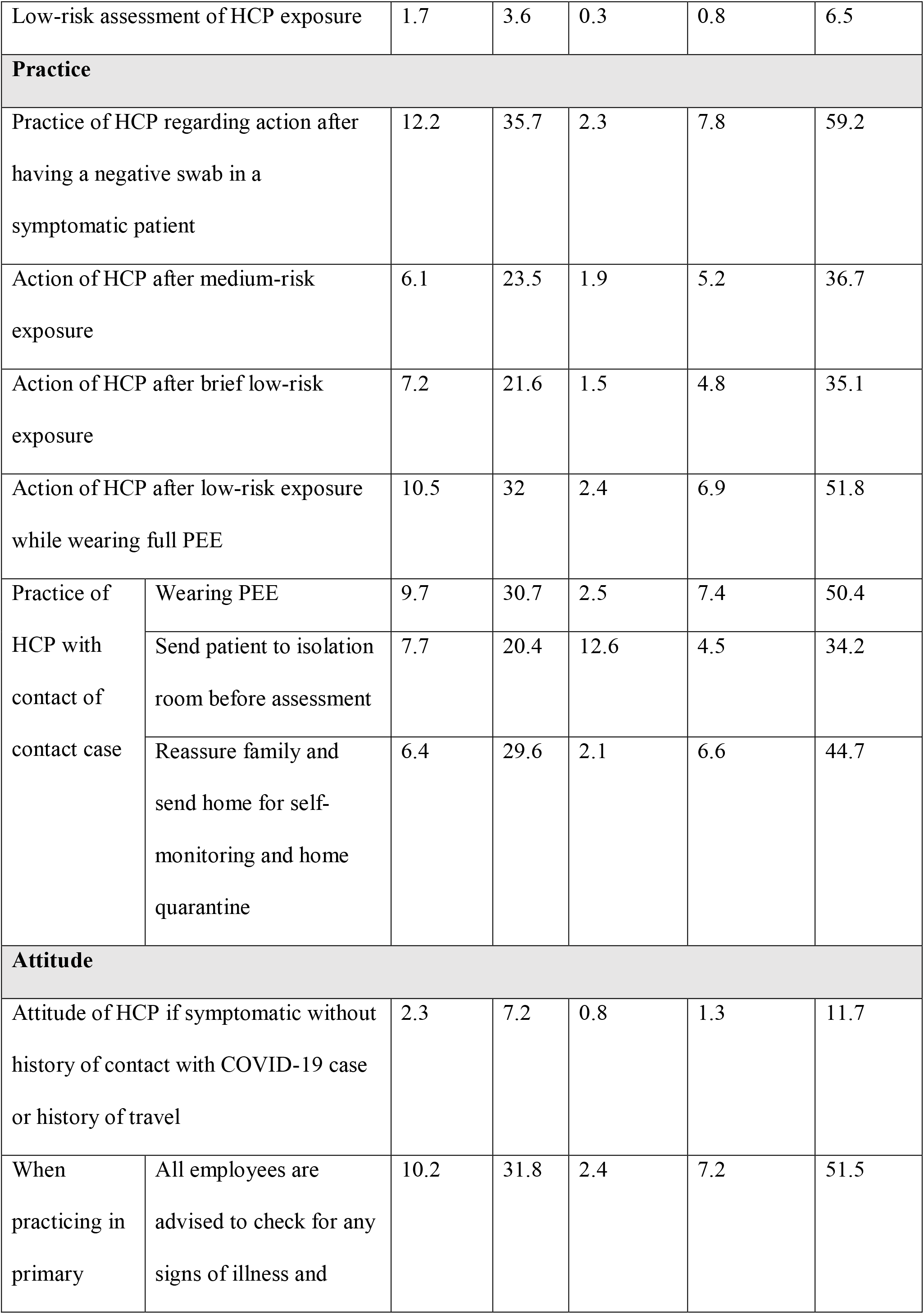
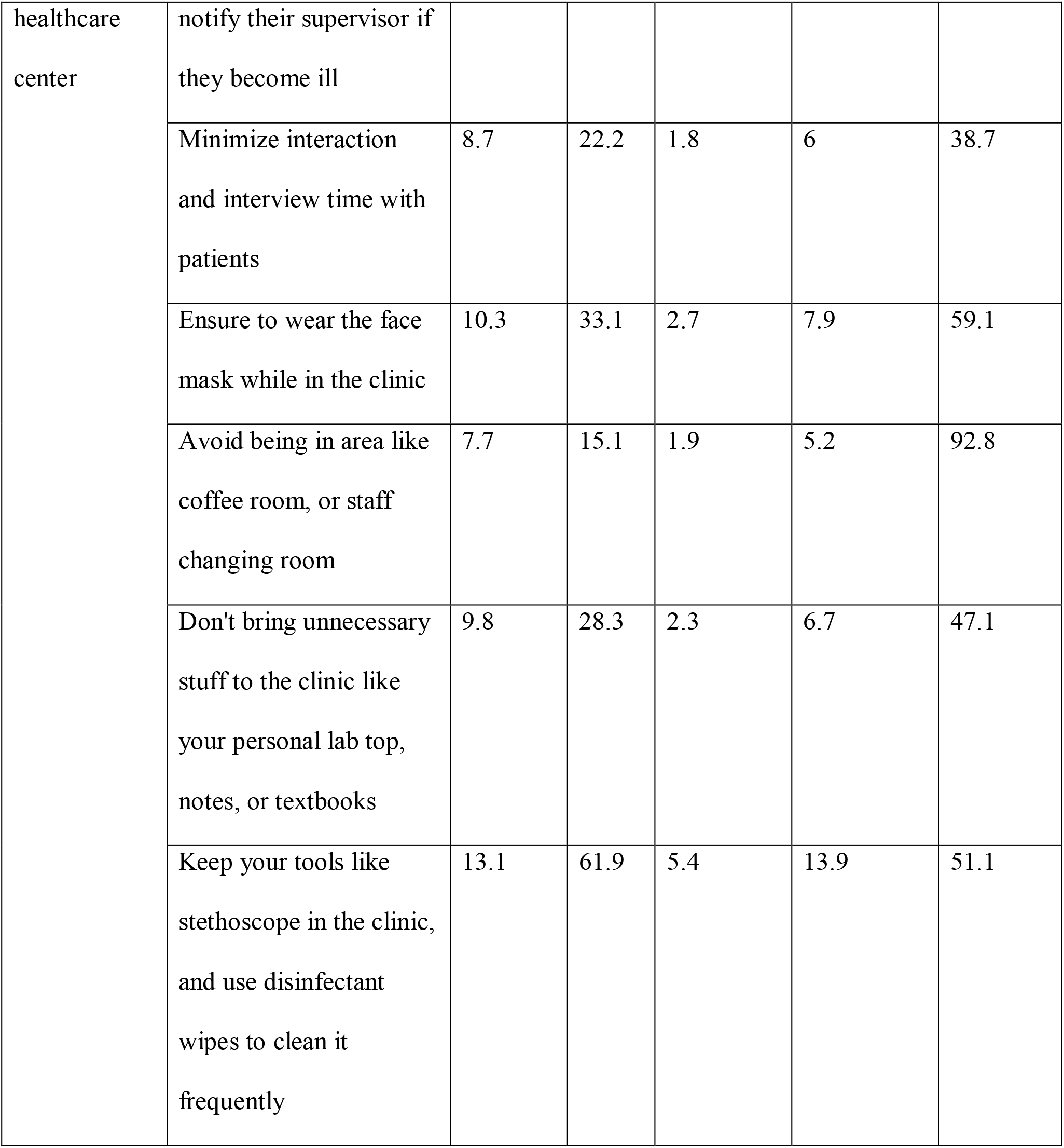
knowledge, practice, and altitude (KAP) of health care providers

Regarding risk assessment, 67.2% of all healthcare providers answered correctly to the question regarding high-risk assessment for a health care provider who was not wearing full PPE when exposed to a positive case of COVID-19, including not wearing a face mask. Nurses did better, with 41.7% answering positively, while only 11.7% of physicians answered positively, whereas the technicians group was significantly positive (P value 0.016). In contrast, high-risk assessment knowledge was significantly negatively associated with consultants and residents (P value was .030 and .037, respectively).

Of healthcare providers’ exposure while performing tooth extraction in a case of COVID-19, while wearing surgical masks and hand gloves with a medium-risk total exposure time of 20 minutes, 30% got the right answer. Of the nurses, 18.5% answered correctly, while only 5.2% of physicians and 4.3% of technicians got the right answered correctly. Knowledge of medium-risk exposure assessment of healthcare providers was significantly and positively associated with the surgical specialty group (P value 0.019). Moreover, medium-risk assessment knowledge was significantly and negatively associated with age (P value was 0.009).

For the question on low-risk exposure assessment, most healthcare providers overestimated the risk of high-risk exposure (52%). Knowledge of low-risk exposure assessment of healthcare providers was significantly and positively associated with the consultant group (P value<0.0001).Table (3)

**Table 3:**
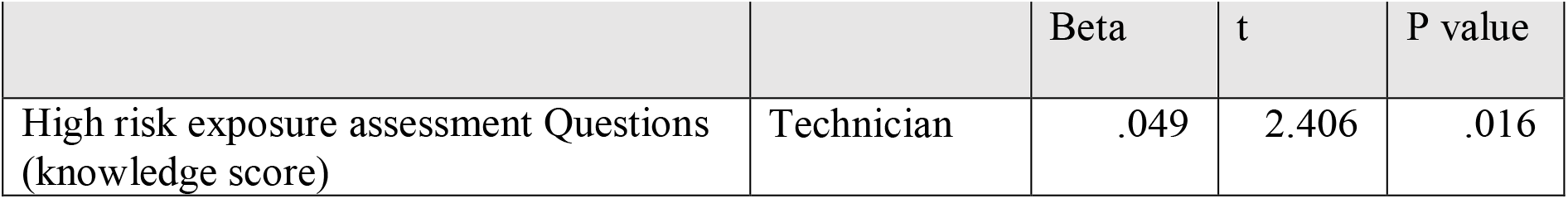
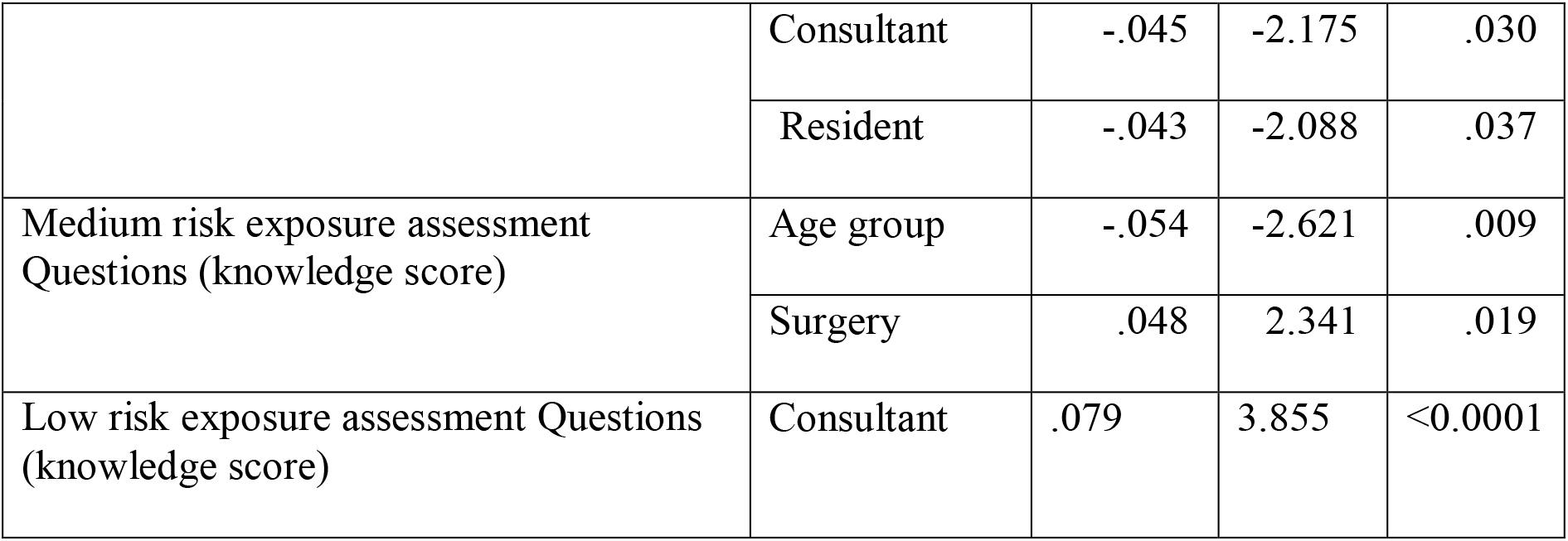
linear regression of healthcare providers knowledge about exposure risk assessment

|  |  | Beta | t | P value |
| --- | --- | --- | --- | --- |
| High risk exposure assessment Questions (knowledge score) | Technician | .049 | 2.406 | .016 |
|  | Consultant | -.045 | -2.175 | .030 |
|  | Resident | -.043 | -2.088 | .037 |
| Medium risk exposure assessment Questions (knowledge score) | Age group | -.054 | -2.621 | .009 |
|  | Surgery | .048 | 2.341 | .019 |
| Low risk exposure assessment Questions (knowledge score) | Consultant | .079 | 3.855 | <0.0001 |

Attitude of healthcare workers toward COVID-19 testing was assessed through the question “what action they will do, if they start to develop any symptoms like dry cough in the absence of a history of contact to COVID-19 case or travel history. Nearly half of them (47.5%) reported that they would ‘‘do a COVID-19 PCR test’’ as an action, while 11.7% would continue to work without further action and 6.3 % would isolate themselves at home. More than half of the HCWs practicing from the primary healthcare clinic reported that they will check for any signs of illness and notify their supervisors if they become ill, ensure wearing a face mask while in the clinic, and keep tools like the stethoscope in the clinic and use disinfectant wipes to clean it frequently; 51.1%,54.1%, and 51.1%). Similarly, 47.1% reported that they would not bring unnecessary staff to the clinic, and only 29.8% reported that they would avoid being in areas like coffee rooms and staff changing rooms. With regard to the COVID-19 practice among healthcare workers, 59.9% of all healthcare providers correctly answered questions related to the action after having a negative swab in a symptomatic patient, of which35.7% were nurses, and only 12.2% were physicians. Medium-risk exposure assessment was correctly answered by 36.7% of all healthcare providers of which 23.5% of nurses answered correctly, with only 6.1% physicians and 5.2% technicians. Low-risk exposure assessment was correctly answered by 35.1% of healthcare providers and 51.8% of all healthcare providers answered correctly regarding low-risk exposure with full PPE.

Dealing with a contact of contact to positive cases was answered incorrectly by 50% of all healthcare providers who reported that they would wear full PEE before dealing with such cases, while only 34.2% reported that they would direct the contact of a positive case to the isolation room. In addition, 44.7% of healthcare providers reported that they would reassure the family and send the case home with self-monitoring and home quarantine.

Only 36.5% of all healthcare providers reported that they are always practicing infection control precautions in primary health care and only 1% reported never practicing infection control precautions.

### Difference in Healthcare Provider Knowledge, Attitude and Practice by Demographics

Overall, more than half of all healthcare providers (55.7%) had good direct knowledge. In practice, 48% had good practices toward COVID-19. The overall attitude mean was 2.8, from a maximum score of 7, indicating a positive attitude toward COVID-19.

Comparing different age groups of healthcare providers, ages 41 to 50 had the best knowledge (59.8%) and practice (51.3%). The P value was significant for good knowledge (0.042%), but insignificant for good practice (0.177%). A higher mean for attitude was noted in the same age group (41-50), which reflects a positive attitude.

Higher knowledge (62.4%) and practice (51.5%) scores and attitude mean (3.48) were noted in the group with more years of experience (more than 30 years) as healthcare providers With and insignificant P value for years of experience.

For occupation, specialists had the highest scores in knowledge (62.2%), practice (53.1%), and attitude mean (3.4), with a significant P value for practice (0.015). On the other hand, residents of all specialties had the lowest score, indicating poor knowledge (55.6%) and poor practice (65.6%).

Among the different specialties, pediatrics had the highest knowledge score (64.8%) with a significant P value (0.018), highest practice score (53.9%) with significant P value (0.039), and the highest means for positive attitude of all specialties (3.37). (Table 4) With linear regression, age group, years of experience, pediatric specialty, and specialist occupation were positively correlated with better knowledge scores. Gender was negatively associated with knowledge scores, indicating that females had better knowledge scores. The P value was significant for pediatric specialty and for specialists.

**Table 4:** Difference in health care provider’s knowledge, practice, and altitude (KAP) by demographics

| Table 4: Difference in health care provider's knowledge, practice, and attitude (KAP) by demographics |  |  |  |  |  |  |  |  |  |
| --- | --- | --- | --- | --- | --- | --- | --- | --- | --- |
| Characteristics | Knowledge* |  |  | Attitude** |  |  | Practice*** |  |  |
|  | Poor knowledge | Good knowledge | P value | Mean | SD | P value | Poor Practice | Good Practice | P value |
|  | (%) | (%) |  |  |  |  | (%) | (%) |  |
| **Gender** |  |  | 0.144 |  |  |  |  |  | 0.108 |
| male | 46.8 | 53.2 |  | 2.85 | 2.68 |  | 54.8 | 45.2 |  |
| female | 43.4 | 56.6 |  | 2.80 | 2.77 |  | 51 | 49 |  |
| **Age group** |  |  | 0.042 |  |  | 0.054 |  |  | 0.177 |
| less than 30 | 48.1 | 51.9 |  | 2.6 | 2.68 |  | 56 | 44 |  |
| 30-40 | 46.3 | 53.7 |  | 2.7 | 2.67 |  | 52.4 | 47.6 |  |
| 41-50 | 40.2 | 59.8 |  | 3.03 | 2.69 |  | 48.7 | 51.3 |  |
| more than 50 | 42.5 | 57.5 |  | 3 | 2.79 |  | 53 | 47 |  |
| **Years of experience** |  |  | 0.055 |  |  | 0.324 |  |  | 0.375 |
| less than 5 | 44.1 | 55.9 |  | 2.8 | 2.7 |  | 55 | 45 |  |
| 6-10 | 48.6 | 51.4 |  | 2.6 | 2.7 |  | 54.1 | 45.9 |  |
| 11-20 | 43.8 | 56.2 |  | 2.8 | 2.66 |  | 50.1 | 49.9 |  |
| 21-30 | 40 | 60 |  | 3.08 | 2.7 |  | 51.6 | 48.4 |  |
| more than 30 | 37.6 | 62.4 |  | 3.48 | 2.86 |  | 48.5 | 51.5 |  |
| **Occupation** |  |  | 0.056 |  |  | 0.00 |  |  | 0.015 |
| consultant | 44.1 | 55.9 |  | 3.03 | 2.86 |  | 55.9 | 44.1 |  |
| Nurse | 44.6 | 55.4 |  | 2.73 | 2.63 |  | 51.1 | 48.9 |  |
| pharmacist | 49.6 | 50.4 |  | 2.62 | 2.77 |  | 60.5 | 39.5 |  |
| Resident | 55.6 | 44.4 |  | 2.23 | 2.76 |  | 65.6 | 34.4 |  |
| Specialist | 37.8 | 62.2 |  | 3.4 | 2.81 |  | 46.9 | 53.1 |  |
| Technician | 43.3 | 56.7 |  | 3.01 | 2.76 |  | 52.1 | 47.9 |  |
| **Specialty** |  |  | 0.018 |  |  | 0.014 |  |  | 0.039 |
| Family medicine | 47 | 53 |  | 2.75 | 2.73 |  | 52.4 | 47.6 |  |
| Internal medicine | 48.8 | 51.2 |  | 2.37 | 2.53 |  | 58 | 42 |  |
| Obstetrics and gynecology | 40.6 | 59.4 |  | 3.05 | 2.6 |  | 46.7 | 53.3 |  |
| Pediatric | 35.2 | 64.8 |  | 3.37 | 2.67 |  | 46.1 | 53.9 |  |
| Psychiatry | 43.6 | 56.4 |  | 2.7 | 2.59 |  | 69.2 | 30.8 |  |
| Surgery | 38.9 | 61.1 |  | 3.0<br>5 | 2.69 |  | 50.5 | 49.5 |  |
| others | 46.2 | 53.8 |  | 2.77 | 2.72 |  | 52.2 | 47.5 |  |
| *knowledge section total score ranges from 0-18 and the cut-off level of <than or equal of 8 was set for poor knowledge and > or equal of 9 for good knowledge.<br>** Attitude section total score ranges from 0-7<br>***Practice items total score ranged as 0-9, and score of <5 indicates poor practice toward precautionary measures of COVID-19. |  |  |  |  |  |  |  |  |  |

Regarding the practice score, resident occupation and age were negatively associated with practice scores. The P value was significant for resident occupation (0.032). Years of experience were positively associated with practice scores, which means experience can improve practice. The practice score was negatively associated with gender, which means that females had better practice scores. The P value was not significant for all the mentioned variables.

Positive attitude toward COVID-19 was positively associated with older age, more years of experience in pediatric specialty, and for specialists. Gender was a significant determinant of attitude, indicating that females had a positive attitude toward COVID-19 compared with males, and the P value was not significant (.353). Another determinant was being a pediatrician and holding a specialist position (P value 0.002 and 0.000, respectively). (Table 5) Depression was negatively associated with the owverall performance scores, which means that those with good knowledge, practice, and positive attitudes had lower rates of depression (P value 0.00).

**Table 5:** Linear regression analysis for factors associated with good knowledge, practice, and altitude in healthcare providers.

|  |  | Beta | t | P value |
| --- | --- | --- | --- | --- |
| Knowledge<br>score | Age group | .023 | .740 | .740 |
|  | Years of experience | .025 | .828 | .828 |
|  | gender | -.031 | -1.438 | -1.438 |
|  | Specialty (Pediatric) | 0.064 | 2.928 | 0.003 |
|  | Occupation(specialist) | 0.044 | 1.958 | 0.05 |
| Practice<br>Score | Age group | -.005 | -.165 | .869 |
|  | Years of experience | .036 | 1.182 | .238 |
|  | gender | -.032 | -1.480 | .139 |
|  | Occupation (Resident) | -0.047 | -2.140 | 0.032 |
| Attitude<br>Score | Age group | .018 | .577 | .564 |
|  | Years of experience | .035 | 1.143 | .253 |
|  | gender | -.020 | -.929 | .353 |
|  | Specialty(Pediatric) | 0.068 | 3.148 | 0.002 |
|  | Occupation(specialist) | 0.087 | 3.873 | 0.000 |

## Discussion

Overall performance score results were disappointing. Only 9.2% scored more than 30 out of 40, which indicates good knowledge. Of the participants, 41.8% had intermediate knowledge scores between and 30-20 out of 40. These findings suggest gaps in knowledge, practice, and attitudes. Poor scores might be related to the fact that the questionnaire was based on case-base scenarios and did not include many direct questions. It was also carried out during the early period of the COVID-19 outbreak in the UAE. Moreover, some questions measured multiple aspects simultaneously. The poor overall performance score was similar to a study carried out in the United Arab Emirates, which reported a significant knowledge gap between the amount of information available and the depth of knowledge about COVID-19 among HCWs. ^6^

It is reassuring that most high-risk assessment situations were identified by most of our healthcare providers. Moreover, low-risk assessment exposure was overestimated as high-and moderate-risk exposures.

A total of 55.7% of healthcare providers had good overall direct knowledge scores regarding different knowledge measures: mode of transmission, protective measures, risk assessment, and test sensitivity and specificity. Females had better knowledge scores than male HCPs. Similar results were noted with practice scores and positive attitudes, which concluded that good knowledge results in better practice and positive attitude. A study done in Greece showed a similar conclusion finding in our study regarding the knowledge, practice, and attitude in relation to HCP gender is still questionable due to insignificant P values.^8^

With age being a key factor, the age group from 41 to 50 years has better knowledge and practice scores and a more positive attitude. The same results were also noted with more years of experience in the medical field, which means that older age groups with more years of experience have the best knowledge and practice scores and positive attitudes.

Specialists had the best scores for knowledge and practice, and positive attitude, while residents had the worst scores for knowledge and practice, highlighting the importance of modifying the postgraduate training to include competencies related to infection control early in the training. Ayinde et al. ^5^ showed that similar findings of occupation were significantly associated with knowledge. Worth noting is that pediatricians had significantly higher scores for knowledge, practice, and attitude mean. This may be because they have respiratory infections in a large part of their practice, although this needs to be explored and implemented in the other specialties.

This study utilized case-based scenarios and successfully highlighted the important gaps and associations. Educational programs in infection control may benefit from a similar approach with stratification of risk and responses tailored to it based on evidence-based recommendations. Simulated real-life case drills are an opportunity best to prepare the healthcare setting for similar infectious disease outbreaks.

## Conclusion

The study-demonstrated gaps in specific aspects of knowledge and practice should be focused on in future education and HCP awareness. A structured training program targeting all HCPs, including physicians, nurses, pharmacists, and technicians, is needed in order to have good clinical knowledge and practice about COVID-19.

## Limitations of the study

The participants in the study were mainly from Abu Dhabi City. This limits the generalizability of the study findings to other UAE emirates, although the respondents were from the public and private sectors. Social desirability bias may have occurred because the questionnaire was self-administered. However, anonymity of the questionnaires was maintained. Despite these limitations, we believe that this study might be a reasonable source of information.

## Data Availability

All data were available without restrictions. All relevant data are presented in the paper .

## Abbreviations

COVID-19: Coronavirus Disease-2019
WHO: World Health Organization
KAPs: knowledge, attitudes and practices
OR: odds ratio;
HCW: healthcare worker
HCPs: healthcare providers

## Availability of data and material

All data were available without restrictions. All relevant data are presented in the paper and supplementary files.

## Authors’ contributions

**AKA, LBA, HMA, and RAA** : conception and design and acquisition of data.

**AKA**, **AAA, LBA** : Analysis and interpretation of data. Drafting the article or substantively revising it critically for important intellectual content.

All authors read and approved the final manuscript.

## Acknowledgements

Not applicable

## Ethics approval and Consent to participate

Ethical clearance was obtained from the Institutional Review Board (IRB). Consent was obtained from all participants. All participants were informed about the study, and their participation was voluntary. The information provided by the participants was anonymous and kept confidential.

